# Examining the Link Between Emergency Department Arrival Mode, Social Determinants of Health, and Social Service Needs in Pediatric Emergency Medicine: A Cross-Sectional Study

**DOI:** 10.1101/2025.09.10.25335528

**Authors:** Spencer C. Buted, Mairead Dillon, Meredith J. Meadows, Carl Elston, Alexandria A. Mahoney, Kathleen M. Adelgais

**Author notes:** **Corresponding Author Information** Spencer C. Buted, University of Colorado School of Medicine, Aurora, CO, USA 13001 E 17th Pl, Aurora, CO, USA 80045.

## Abstract

**Background:** Unmet social needs can increase the risk of adverse health outcomes in children. Emergency medical services (EMS) utilization is higher among families facing challenges with certain social determinants of health (SDoH). Understanding service needs among families seeking emergency care may inform interventions designed to support better health outcomes.

**Methods:** We conducted a cross-sectional survey of caregivers and EMS clinicians presenting to a pediatric emergency department (ED) via EMS or private vehicle (POV). Participants completed a survey reporting access to transportation, food, housing, healthcare, mental healthcare, and financial support. We evaluated differences in self-reported SDoH and service needs by arrival mode and differences in caregiver-reported needs with those perceived by EMS clinicians using Pearson’s chi-squared and Fisher’s exact tests.

**Results:** A total of 159 caregivers (111 POV, 48 EMS) and 17 EMS clinicians were enrolled. Caregiver demographics were similar between groups. Overall, 67 (42%) caregivers reported at least one service need with few differences between groups. Difficulty seeking healthcare, childcare, and mental health support were most frequently reported. EMS arrivals were more likely to report difficulty seeking child healthcare (p=.02); POV arrivals were more likely to report personal mental health needs (p<.01). There was no difference in requests for child mental health treatment (p>.90).

**Conclusions:** Service needs are common in pediatric emergency care. Difficulty accessing child healthcare and mental healthcare were common in our population. Screening pediatric caregivers during emergency care including during EMS activations may identify families in need of support services.

**KEY MESSAGE STATEMENT:** Social Determinants of Health (SDoH) have important health implications in children. In pediatric emergency care, caregivers and children arriving via Emergency Medical Services (EMS) are more likely to have social needs. There is a paucity of research on the specific needs of this population compared to private vehicle arrives and if EMS clinicians are aware of these needs. This study compares the SDoH of caregivers in the ED between arrival type and assesses if EMS clinicians recognize the needs of the people they transport. These results may be useful in improving EMS clinician curricula and addressing SDoH in families with young children.

## INTRODUCTION

Emergency Medical Services (EMS) is a critical component of the healthcare system, positioned at the intersection of public health, public safety, and acute care. EMS clinicians frequently encounter families in their home environments, providing a rare and valuable perspective on the social determinants of health (SDoH)—factors such as housing, food availability, transportation, and access to healthcare—that may be less visible to clinic- or hospital-based providers. These determinants significantly influence health outcomes and healthcare utilization.(1–5)

EMS utilization is disproportionately high among populations with elevated social risk.(2, 5–10) Pediatric EMS encounters are more frequent among families from neighborhoods with lower Child Opportunity Index (COI) scores, reflecting systemic inequities that limit access to preventive care and other supportive services.(11) Notably, between 12–30% of pediatric EMS calls result in non-transport to the emergency department, highlighting EMS as a potential yet underused touchpoint for identifying and supporting families outside of conventional care settings.(12–14) Given their ability to reach nearly all U.S. households, EMS systems may offer an equitable and scalable means of connecting high-risk families with community resources and prior research demonstrates that families are receptive to EMS-facilitated referrals for social support services, yet standardized mechanisms for doing so remain limited.(2, 15)

While EMS may have potential in supporting patients with chronic conditions such as asthma and mental health disorders, less is known about its role in identifying social needs among children and their families.(16) There are only a few reports comparing caregiver reported service needs between those transported by EMS and those who arrive by provide vehicle. Moreover, few studies have explored whether EMS clinicians are able to identify the SDoH needs, and none have examined the extent to which their assessments align with caregiver self-report. (2, 17, 18)

Future interventions that may leverage EMS as a community partner to address upstream drivers of pediatric health require a better understanding of the service needs and social determinants of health among families presenting for emergency care. The primary aim of this study is to compare the caregiver reported social determinants of health and unmet service needs for those arriving to a pediatric emergency department via EMS to those arriving by private vehicle. Secondarily, we sought to examine agreement between EMS clinician’s perception of a family’s service needs and those reported by caregivers.

## METHODS

### Design

We conducted a pilot cross-sectional study utilizing an electronically administered confidential survey. The location of the study was in the emergency department of large, free-standing pediatric hospital.

### Study Population and Participant Involvement

Participants were caregivers of patients ages 0-5 years presenting to the emergency department by EMS transport or by private vehicle (POV). We excluded families arriving by EMS from another medical facility, those presenting with critical illness, or those with clinical concerns for child maltreatment. Eligible caregivers were at least 18 years of age, responsible for the child’s care at least 50% of the time, and literate in English or Spanish. To reduce potential bias, no other information (caregiver age, address, insurance) was used to screen potential participants. The inclusion of multiple languages was in effort to reduce potential bias or confounders.

Emergency Department (ED) arrivals were screened by trained research personnel and approached when not actively receiving care from the medical team. If a caregiver was eligible and arrived via EMS transport, the EMS clinicians transporting the child of the caregiver were considered eligible. Participation was voluntary; all sensitive questions were optional, and participants could withdraw at any time. All responses remained confidential and were stored on a secure server. Given the sensitive nature of certain questions, no identifying information was collected. Participants were involved in this research project. Pilot participants were used to gain insight on study protocols, survey language, and appropriate time to approach. Prior to beginning enrollment, the study was reviewed and approved by the local Institutional Review Board.

### Data Collection

Self -reported caregiver demographic data collected included: age, race, ethnicity, marital status, household income, education level, employment status, and the number of adults in the home. Self-reported EMS demographic data included: race, ethnicity, highest certification level, and years of experience.

Caregivers completed a 19-question composite survey adapted from the iScreen study and the USDA Food Security Module, tools previously utilized in the literature (19, 20) Survey questions specifically inquired about SDoH and service needs including transportation access, food insecurity, housing insecurity, financial insecurity, healthcare access, and mental health needs. We assessed caregivers’ willingness to accept services on a 5-point Likert scale ranging from 1 = Very Unlikely to 5 = Very Likely. Preferred method of connection with services was also collected. If a caregiver indicated a desire to learn more about social services to address their needs during the index ED visit, research personnel notified the primary medical team to connect them with social work.

EMS clinicians completed an accompanying survey assessing their perceptions of the caregiver and family SDoH and service needs just transported. Questions on the EMS survey were matched to the caregiver questions to allow for comparison between the two groups within the EMS-caregiver dyad. EMS clinicians were not approached until after patient handoff to ED personnel to ensure continuity of care. All responses were confidential and voluntary, no information that could identify the clinician or EMS transport service was collected. The enrollment period was over an 18-month period from September 2023 to February 2025.

### Outcome Measures

The primary outcome measure is the proportion of families who report unfavorable SDoH and social service needs correlated with the mode of arrival to the ED. Secondary outcomes include the best method for contact and degree of agreement between EMS perception and family report of SDoH and social service needs.

### Statistical Analysis

Caregiver demographics and survey results were summarized by arrival type, EMS or POV arrival, using counts and percentages for categorical variables and medians and interquartile values for continuous variables. The sample size for the study is based on a power of 80% and a p-value *<* .05 with two-sided chi-square test. Demographics and survey results were compared by arrival type using Fisher’s exact and Pearson’s chi-squared tests for categorical variables and Wilcoxon rank sum tests for continuous variables. P-values < .05 were considered statistically significant.

To compare self-reported social determinants of health and service needs with those reported by EMS clinicians for subjects who arrived by EMS vehicle and had an EMS survey completed, two by two tables were created. Percent agreements between self-reported and EMS reported social determinants of health and social needs were calculated. All statistical analyses were completed using R Statistical Software (version 4.4.1; R Core Team 2024). This manuscript reports methods and findings according to STROBE guidelines.

## RESULTS

During the study period, a total of 160 caregivers and 17 EMS clinicians were enrolled with 48 caregiver respondents arriving by EMS, 111 caregiver respondents arriving by POV. One caregiver decided to not participate following EMS enrollment resulting in 16 caregiver-EMS clinician dyads. Demographics of the caregiver respondents are demonstrated in Table 1.

**Table 1.** Participant Demographics by Arrival Type.

| Characteristic | EMS<sup>1</sup> Arrival<br>N = 48<sup>3</sup> | POV<sup>2</sup> Arrival<br>N = 111<sup>3</sup> | p-value<sup>4</sup> |
| --- | --- | --- | --- |
| <u>Caregiver Relationship</u> |  |  | .90 |
| Mother | 38 (79%) | 83 (75%) |  |
| Father | 8 (17%) | 21 (19%) |  |
| Other | 2 (4.2%) | 7 (6.3%) |  |
| <u>Caregiver Marital Status</u> |  |  | .40 |
| Married | 30 (63%) | 65 (59%) |  |
| Divorced | 3 (6.3%) | 5 (4.5%) |  |
| Widowed | 1 (2.1%) | 0 (0%) |  |
| Never Married | 14 (29%) | 41 (37%) |  |
| <u>Household Income</u> |  |  | .30 |
| Less than \$20,000 | 9 (19%) | 16 (15%) | |
| \$20,000 - \$34,999 | 9 (19%) | 14 (13%) | |
| \$35,000 - \$49,999 | 3 (6.4%) | 19 (17%) | |
| \$50,000 - \$74,999 | 6 (13%) | 22 (20%) | |
| \$75,000 - \$99,999 | 6 (13%) | 11 (10%) | |
| \$100,000+ | 14 (30%) | 27 (25%) | |
| <u>Caregiver Education Level</u> |  |  | .07 |
| Some high school | 9 (19%) | 7 (6.4%) |  |
| High school diploma or equivalent | 12 (25%) | 19 (17%) |  |
| Vocational training certificate or equivalent | 0 (0%) | 6 (5.5%) |  |
| Some college | 9 (19%) | 30 (27%) |  |
| College degree | 13 (27%) | 26 (24%) |  |
| Master's degree | 5 (10%) | 18 (16%) |  |
| Doctoral degree | 0 (0%) | 4 (3.6%) |  |
| <u>Caregiver Employment Status</u> |  |  | .70 |
| Full-time | 22 (46%) | 55 (50%) |  |
| Part-time | 10 (21%) | 13 (12%) |  |
| Unemployed and looking for work | 2 (4.2%) | 7 (6.4%) |  |
| Unemployed | 7 (15%) | 10 (9.1%) |  |
| Student | 1 (2.1%) | 5 (4.5%) |  |
| Retired | 0 (0%) | 1 (0.9%) |  |
| Homemaker | 5 (10%) | 9 (8.2%) |  |
| Self-employed | 1 (2.1%) | 7 (6.4%) |  |
| Unable to work | 0 (0%) | 3 (2.7%) |  |
| <u>Caregiver Race</u> |  |  | .50 |
| White | 26 (55%) | 63 (57%) |  |
| Black | 5 (11%) | 17 (15%) |  |
| Asian | 3 (6.4%) | 6 (5.5%) |  |
| Native American, Native Alaskan | 1 (2.1%) | 2 (1.8%) |  |
| Pacific Islander | 1 (2.1%) | 0 (0%) |  |
| More than One Race | 1 (2.1%) | 6 (5.5%) |  |
| Other | 9 (19%) | 11 (10%) |  |
| Prefer Not to Answer | 1 (2.1%) | 5 (4.5%) |  |
| <u>Caregiver Ethnicity</u> |  |  | .70 |
| Hispanic or Latino | 17 (35%) | 32 (29%) |  |
| Not Hispanic or Latino | 29 (60%) | 73 (66%) |  |
| Prefer Not to Answer/No Response | 2 (4.2%) | 6 (5.4%) |  |
| Number of Adults in Child's Home | 2.00 (2.00, 2.00) | 2.00 (2.00, 2.00) | >.90 |
| Number of Children in Home | 2.00 (1.00, 3.00) | 2.00 (1.00, 3.00) | .12 |
| <sup>1</sup>Emergency Medical Services |  |  |  |
| <sup>2</sup>Private Vehicle |  |  |  |
| <sup>3</sup>n (%); Median (Q1, Q3) |  |  |  |
| <sup>4</sup>Fisher's exact test; Pearson's Chi-squared test; Wilcoxon rank sum test |  |  |  |

There were no significant demographic differences between caregivers of subjects who arrived via POV and those who arrived via EMS. EMS clinician demographic data is shown in Supplemental Table 1.

Table 2 displays the caregiver reported SDoH and service needs in the domain of healthcare access.

**Table 2.** Self-Reported Caregiver Healthcare Access and Needs.

| Characteristic | EMS<sup>1</sup> N<br>= 48<sup>3</sup> | POV<sup>2</sup> N =<br>111<sup>3</sup> | p-value<sup>4</sup> |
| --- | --- | --- | --- |
| Has your child ever had a chronic medical condition? (ex. asthma, diabetes, seizures)? | 11 (23%) | 31 (28%) | .50 |
| Do you have concerns about seeking healthcare for your child when they needed it? | 10 (21%) | 9 (8.3%) | .02 |
| Have you had concerns about finding affordable or reliable child care? | 11 (23%) | 26 (24%) | .90 |
| Have you felt at any time that you might need treatment for a mental health problem? | 0 (0%) | 18 (17%) | **.003** |
| Have you felt at any time that another adult in the home might need treatment for a mental health problem? | 1 (2.1%) | 11 (10%) | .11 |
| Have you felt at any time that you might need treatment for an alcohol or drug problem? | 1 (2.1%) | 2 (1.8%) | >.90 |
| Have you felt at any time that another adult in the home might need treatment for an alcohol or drug problem? | 0 (0%) | 2 (1.8%) | >.90 |
| Did you have concerns about adults in the home not seeking healthcare due to lack of insurance or inadequate health insurance? | 5 (10%) | 12 (11%) | .90 |
| Has an adult in the child's household had a chronic medical condition? (ex. asthma, diabetes, seizures) | 5 (11%) | 25 (23%) | .07 |
| <sup>1</sup>Emergency Medical Services |  |  |  |
| <sup>2</sup>Private Vehicle |  |  |  |
| <sup>3</sup>n (%); Median (Q1, Q3) |  |  |  |
| <sup>4</sup>Fisher's exact test; Pearson's Chi-squared test; Wilcoxon rank sum test |  |  |  |

Caregivers arriving by EMS and POV most often reported concerns finding affordable childcare. When comparing caregiver response by arrival type, EMS arrivals were significantly more likely to report concern seeking healthcare for their child when they needed it. POV arrivals were significantly more likely to report that they may personally need treatment for a mental health problem. Caregiver responses to service needs questions are demonstrated in Table 3.

**Table 3.** Caregiver Self-Reported General Service Needs.

| Service Need Type | EMS<sup>1</sup><br>N=48<sup>3</sup> | POV<sup>2</sup><br>N=111<sup>3</sup> | p-value<sup>4</sup> |
| --- | --- | --- | --- |
| Participant responded yes to at least 1 service need | 23 (48%) | 44 (40%) |  |
| Food Assistance (SNAP, WIC, food pantries, etc.) | 7 (15%) | 15 (14%) | .90 |
| Housing Assistance (Section 8 applications, utility stipends, etc.) | 7 (15%) | 9 (8.1%) | .3- |
| Employment Services (career advising, application assistance, etc.) | 4 (8.3%) | 9 (8.1%) | >.90 |
| Childcare Assistance | 1 (2.1%) | 7 (6.3%) | .40 |
| Education Assistance (tutoring, academic advising, admissions help, etc.) | 5 (10%) | 5 (4.5%) | .20 |
| Caregiver health (Medicaid applications, prescription coverage, etc.) | 2 (4.2%) | 1 (0.9%) | .20 |
| Family violence | 1 (2.1%) | 1 (0.9%) | .50 |
| Assistance with public benefits | 7 (15%) | 9 (8.1%) | .30 |
| Mental health care resources for self or child | 4 (8.3%) | 11 (9.9%) | >.90 |
| Child health (Medicaid for children, finding a PCP, etc.) | 8 (17%) | 6 (5.4%) | **.03** |
| Financial assistance | 5 (10%) | 5 (4.5%) | .20 |
| Transportation (public transport funds, affording insurance, etc.) | 3 (6.3%) | 5 (4.5%) | .70 |
| Substance use care | 1 (2.1%) | 1 (0.9%) | .50 |
| <sup>1</sup>Emergency Medical Services |  |  |  |
| <sup>2</sup>Private Vehicle |  |  |  |
| <sup>3</sup>n (%); Median (Q1, Q3) |  |  |  |
| <sup>4</sup>Fisher's exact test; Pearson's Chi-squared test; Wilcoxon rank sum test |  |  |  |

Overall, 42% of families reported at least one service need. Assistance with child health was significantly different between arrival type groups; EMS arrivals were more likely to indicate a desire for child healthcare services. There was no significant difference between arrival groups for mental health service needs. Top service needs reported by caregivers arriving via EMS were child health assistance followed by food assistance, housing assistance, and help with enrollment in public benefits. Top service needs reported by caregivers arriving via POV were food assistance and mental health care. Overall, 5 caregivers requested to be connected with social work at the emergency department. Of those, 4 caregivers arrived via EMS transport and 1 arrived via POV.

Dyad responses include arrivals where both the caregiver and transporting EMS clinician completed their respective survey tools. Figure 1 reports the percent agreement in survey results of complete dyads. Five SDoH and service needs had 100% concordance with the lowest percent agreement at 65% for access to affordable childcare. All other responses had percent agreements greater than 80%.

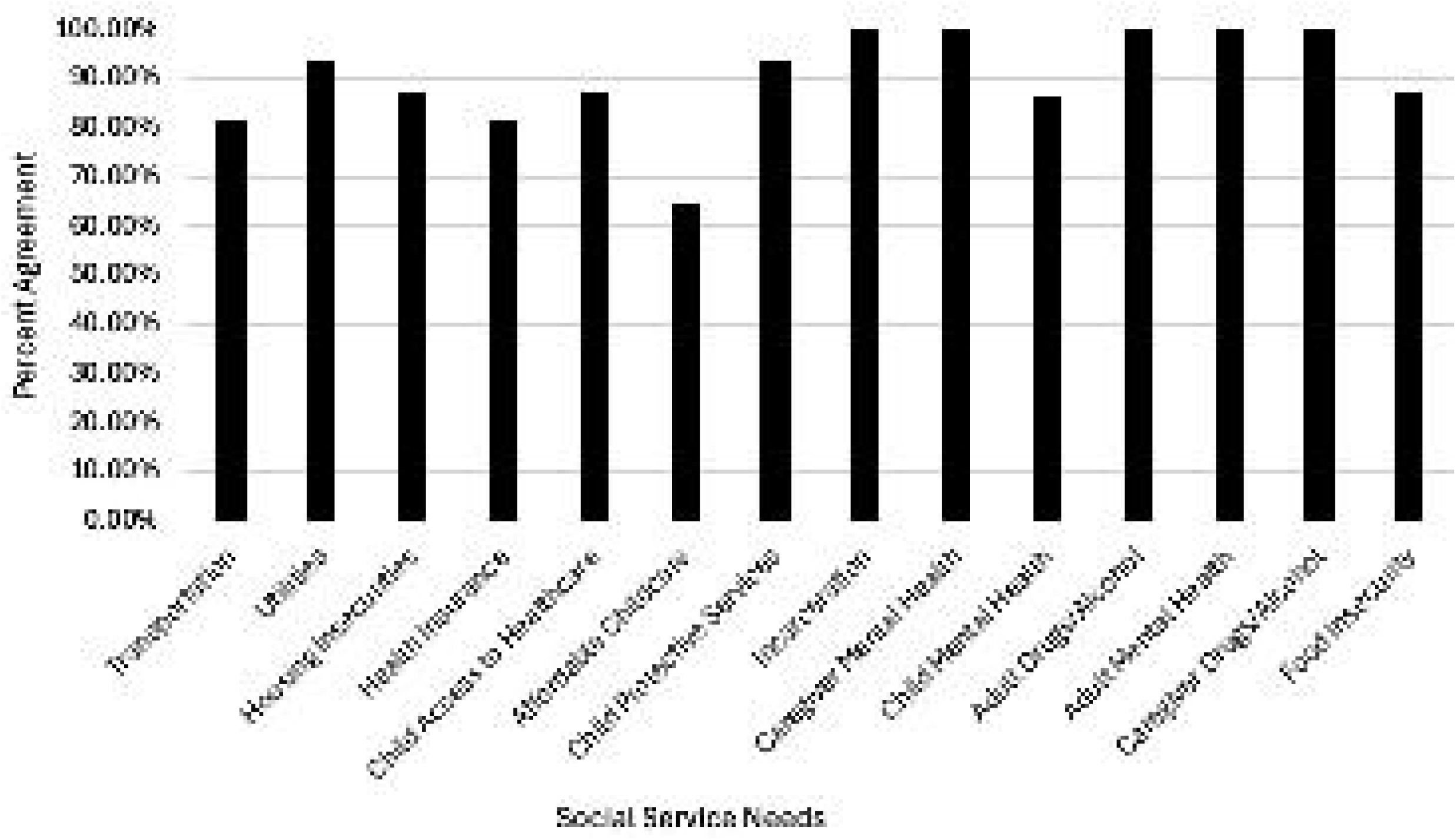

## DISCUSSION

EMS clinicians frequently serve high-risk pediatric populations but are underutilized in addressing social determinants of health. Little is known about how the perceptions of EMS clinicians align with caregiver reports of service needs or how the needs of families arriving via EMS differ from those arriving by POV. The primary aim of this study was to determine the differences in SDoH and service needs between families arriving to the emergency department by POV and families arriving by EMS transport.

In contrast to prior studies, we did not find an increased need for transportation among families transported by EMS.(2) Several factors may explain the difference in reported service needs in our study compared to prior research. First, differences in screening tools or the phrasing of questions can significantly influence how families interpret and respond to items. It is also possible that transportation access is not a problem, and the activation of EMS was related to caregiver concerns of illness severity in their child. Additionally, regional and population-level differences may play a role; our hospital may serve a community with better access to transportation.

Similar to prior studies, however, we found the most common service needs reported by caregivers are trouble seeking child healthcare, finding childcare, and needing mental health support, with no differences based on arrival mode. (16, 18) Concern with seeking healthcare can stem from a broad range of underlying social determinants including distance to care, insurance coverage, financial status, or low medical literacy.(2, 18, 19) With higher rates of EMS utilization, these caregivers may be calling EMS for any concern with their child.(2, 5, 6) This makes EMS systems a potential tool to identify these barriers and connect families with services to address these unmet service needs.

Following the COVID-19 pandemic, the burden of mental health conditions – including self-harm and anxiety disorders significantly increased across the United States. (21) Caregivers bringing their child to the ED were significantly more likely to report a need for treatment for their own mental health problems. While the primary patient is the child, targeted interventions to caregivers’ mental health care may be a way to provide necessary support to the family and reduce risk of adverse childhood experiences (ACEs). (22–24) Emergency department staff may be able to identify and support families with at-risk children, mitigating the potential negative impact of untreated mental health problems.

A secondary aim of this study was to evaluate the extent to which of EMS clinicians could detect service needs of the families that they transport to the ED. To our knowledge, no other studies have assessed the concordance of EMS detection of SDoH and service needs with self-reported needs in a population of families seeking care for a child. We hypothesized that EMS would be able to accurately report social services needs of the families they transport. In our small sample, there was high concordance between caregiver responses and EMS clinician responses with strong concordance in dyads in reporting mental health needs and substance use in the home.

Although there is high concordance in survey responses, several studies have shown that EMS documentation of SDoH is rare and inconsistent.(9, 25) The implementation of standardized documentation practices may improve visibility of valuable EMS clinician insight into family needs.

The most frequent domain with discordance was the caregiver experiencing trouble with finding affordable childcare. This is not surprising because understanding child care needs require an understanding of the caregiver’s life situation, and it would be difficult for EMS clinicians to gain this insight in an encounter where there is a child with an acute illness. Of note, our overall sample size was small, and future research is required however, our study highlights an opportunity to explore the role of screening practices with EMS clinician and provides a framework for future investigation.

Importantly, community paramedicine is an emerging model of care in which specially trained EMS professionals provide non-emergent, preventative, and follow-up services in community settings. These programs aim to reduce avoidable emergency department visits and hospitalizations by addressing patients’ medical, behavioral, and social needs where they live. (26) Families who call EMS for their children are more likely to experience challenges accessing timely, consistent, and preventive pediatric healthcare—often due to unmet social needs.(5, 6) Application of community paramedicine programs to support families may be a valuable resource, as it offers a proactive, home-based approach that may address the upstream factors that contribute to health disparities.(27) Our results can help inform future community paramedicine research and outcomes. The study has generalizability by addressing common and widespread SDoH among caregivers and leveraging EMS clinician perspectives, which are relevant across diverse healthcare settings. EMS personnel interact with patients and their caregivers in unique ways. They may witness family dynamics in the household; this gives them access to information not available to personnel within the emergency department. Among our small group of EMS-caregiver dyads, we did fine that EMS clinicians detected the social needs of the families that they transport with high rates of concordance compared to self-reported needs. Future EMS clinician training curricula may benefit from the addition of modules focused on the social needs of families and the impact of SDoH on health outcomes.

### Limitations

The subjects in this study represent a convenience sample of caregivers, limiting the generalizability and external validity of the results. This study was conducted at a free-standing, quaternary pediatric hospital which may limit the generalizability of these results. Selection bias may also have contributed, as families experiencing the highest levels of social risk may have been underrepresented due to exclusion. Social needs and social determinants of health were self-reported, so these responses may have been affected by recall bias. Respondents may also have experienced social desirability bias, because of the sensitive nature of the survey, leading them to underreport true social service needs. We attempted to mitigate this bias by keeping participant responses confidential and anonymous. Finally, the timing and context of an EMS encounter—often occurring during a moment of acute stress—may limit a caregiver’s ability or willingness to fully engage in screening or disclose non-urgent service needs.

### Conclusions

The findings of this study highlight that mode of arrival is associated with distinct SDoH and social needs among caregivers of presenting to a pediatric emergency department. Caregivers arriving via EMS frequently reported difficulty accessing healthcare for their children, suggesting that interventions such as community paramedicine programs may help reduce EMS utilization rates while improving overall public health outcomes. There was a high level of concordance between caregivers’ self-reported service needs and those identified by EMS clinicians, underscoring the potential value of EMS personnel in recognizing and addressing unmet social needs in pediatric populations.

## Data Availability

All data produced in the present study are available upon reasonable request to the authors

## AUTHOR CONTRIBUTORSHIP

**Spencer C. Buted:** Conceptualization (equal), Methodology (equal), Formal Analysis (supporting), Investigation (lead), Resources (lead), Data Curation (lead), Writing – Original Draft (lead), Visualization (equal), Project Administration (equal) **Mairead Dillon:** Conceptualization (supporting), Methodology (supporting), Software (lead), Validation (equal), Formal Analysis (lead), Data Curation (supporting), Writing – Review and Editing (supporting), Visualization (equal) **Meredith J. Meadows:** Investigation (supporting), Writing – Original Draft (supporting) **Carl Elston:** Methodology (supporting), Investigation (supporting), Resources (supporting), Project Administration (supporting) **Alexandria A. Mahoney:** Investigation (supporting) **Kathleen M Adelgais:** Conceptualization (equal), Methodology (equal), Formal Analysis (supporting), Resources (supporting), Data Curation (supporting), Writing – Review and Editing (lead), Visualization (equal), Supervision (lead), Project Administration (equal).

## ACKNOWLEDGEMENTS

None

