## Supplemental Table 1 for "Examining the Link Between Emergency Department Arrival Mode, Social Determinants of Health, and Social Service Needs in Pediatric Emergency Medicine: A Cross-Sectional Study"

**SUPPLEMENTAL MATERIALS**

| **Characteristic** | **N = 17***^1^* |
| --- | --- |
| Race |  |
| White | 12 (71%) |
| Black | 0 (0%) |
| Asian | 1 (5.9%) |
| Native American, Native Alaskan | 0 (0%) |
| Pacific Islander | 1 (5.9%) |
| More than One Race | 1 (5.9%) |
| Other | 1 (5.9%) |
| Prefer Not to Answer | 1 (5.9%) |
| Ethnicity |  |
| Hispanic or Latino | 3 (18%) |
| Not Hispanic or Latino | 13 (76%) |
| Prefer Not to Answer | 1 (5.9%) |
| Highest Certification Level |  |
| EMT | 7 (41%) |
| AEMT | 0 (0%) |
| Paramedic | 10 (59%) |
| Other | 0 (0%) |
| Years of Practice | 4 (2, 6) |
| *^1^*n( %); Median (Q1, Q3) |  |

*Supplemental Table 1.* EMS Clinician Demographics
